# Kinetics of naturally induced binding and neutralizing anti-SARS-CoV-2 antibody levels and potencies among Kenyan patients with diverse grades of COVID-19 severity

**DOI:** 10.1101/2022.10.05.22280716

**Authors:** John Kimotho, Yiakon Sein, Shahin Sayed, Reena Shah, Kennedy Mwai, Mansoor Saleh, Perpetual Wanjiku, Jedidah Mwacharo, James Nyagwange, Henry Karanja, Bernadette Kutima, John Gitonga, Daisy Mugo, Ann Karanu, Linda Moranga, Vivian Oluoch, Jasmit Shah, Julius Mutiso, Alfred Mburu, Zaitun Nneka, Peter Betti, Wanzila Usyu Mutinda, Abdirahman Issak Abdi, Philip Bejon, Lynette Isabella Ochola-Oyier, George M. Warimwe, Eunice Nduati, Francis M. Ndungu

## Abstract

**Background:** Given the low levels of COVID-19 vaccine coverage in Sub-Saharan Africa, despite high levels of natural SARS-CoV-2 exposures, strategies for extending the breadth and longevity of naturally acquired immunity are warranted. Designing such strategies will require a good understanding of natural immunity.

**Methods:** We used ELISA to measure whole-spike IgG and spike-receptor binding domain (RBD) total immunoglobulins (Igs) on 585 plasma samples collected longitudinally over five successive time points within six months of COVID-19 diagnosis in 309 COVID-19 patients. We measured antibody neutralizing potency against the wild-type (Wuhan) SARS-CoV-2 pseudo-virus in a subset of 51 patients over three successive time points. Binding and neutralizing antibody levels and potencies were then tested for correlations with COVID-19 severities, graded according to the National Institute of Health (NIH), USA criteria.

**Results:** Rates of sero-conversion increased from Day 0 (day of PCR testing) to Day 180 (six months) (63.6% to 100 %) and (69.3 % to 97%) for anti-spike IgG and anti-spike-RBD binding Igs, respectively. Levels of these binding antibodies peaked at Day 28 (P<0.0001) and were subsequently maintained for six months without significant decay (p>0.99). Similarly, antibody neutralizing potencies peaked at Day 28 (p<0.0001) but had decreased by three-folds, six months after COVID-19 diagnosis (p<0.0001). Binding antibodies levels were highly correlated with neutralizing antibody potencies at all the time points analyzed (r>0.6, P<0.0001). Levels and potencies of binding and neutralizing antibodies increased with disease severity.

**Conclusion:** Most COVID-19 patients from Sub-Saharan Africa generate SARS-CoV-2 specific binding antibodies that remain stable during the first six months of infection. Although antibody binding levels and neutralizing potencies were directly correlated, the respective neutralizing antibodies decayed three-fold by the sixth month of COVID-19 diagnosis suggesting that they are short-lived, consistent with what has been observed elsewhere. Thus, just like for other populations, regular vaccination boosters will be required to broaden and sustain the high levels of predominantly naturally acquired anti-SARS-CoV-2 neutralizing antibodies.

## Introduction

Our understanding of the nature and role of anti-SARS-CoV-2 (severe acute respiratory syndrome coronavirus-2) antibody responses in immunity to COVID-19 (corona virus disease-2019) is only beginning to be realized now. Antibodies, both infection and vaccine induced, are the most well established correlate of protection against most of the medically important pathogens (1,2). For example, an anti-hemagglutinin inhibition antibody titer of 1:40 is an established correlate of protection against influenza following vaccination with inactivated influenza vaccine in children and adults (3,4). Antibodies offer protection against viruses through a myriad of mechanisms such as neutralization, opsonization, formation of immune complexes, complement deposition and antibody dependent cellular cytotoxicity (5–7). SARS-CoV-2 virus infection elicits robust IgG, IgM, and IgA antibody responses targeting various epitopes of the virus (8–11). However, only a subset of these elicited antibodies have neutralization function (10,12,13).

Neutralizing antibodies offer protection by binding to viral epitopes, thereby interfering with virus attachment to the host cell receptor (14,15). The receptor binding domain (RBD), which is part of the SARS-CoV-2 spike protein, has been shown to be the target of >90% of neutralizing antibodies, which provides protection by blocking the virus from fusing with angiotensin converting enzyme (ACE2) of host cells (10).

Some of the previous studies in high income countries (Europe, USA) have shown that naturally induced anti-SARS-CoV-2 antibodies, could be short-lived (16–19) while others reported the contrary (20–22). In addition, some COVID-19 patients from these countries have been shown to remain sero-negative for SARS-CoV-2 antibodies despite being positive by RT-PCR (real time-reverse transcription-polymerase chain reaction) (23,24). Together, such data suggests the existence of inter-population differences in people’s abilities to generate and maintain anti-SARS-CoV-2 antibodies. However, there is a paucity of data on the kinetics and functions of anti-SARS-CoV-2 antibodies from Sub-Saharan African populations (25–27).

COVID-19 vaccine coverage in the Sub-Saharan Africa remains very low (28–30). For example in Kenya, approximately 70% of the adult Kenyan population remains unvaccinated with at least two doses, as of September 2022 (31,32). This then provides a unique window to understand naturally acquired immunity to SARS-CoV-2 in our setting, since the majority have acquired immunity through natural infection rather than vaccination (26,33–37). Comparing anti-SARS-CoV-2 immune responses between patients with different clinical phenotypes and determining the presence and longevity of antibody neutralization function, may also begin to provide clues for potential correlates of COVID-19 disease severity. This is critical, given that there has been significantly less severe disease and associated deaths in Sub-Saharan Africa compared to Europe and North America (38–41).

Here, we describe the kinetics of naturally acquired anti-SARS-CoV-2 binding and virus neutralizing antibodies during the first six months of COVID-19 infection in a cohort of Kenyan patients with varying degrees of disease severity. To the best of our knowledge, this is the first longitudinal study evaluating the kinetics of naturally acquired anti–SARS-CoV-2 antibodies for a period that goes up to 6 months post-COVID-19 diagnosis, in a Sub-Saharan African population.

## MATERIALS AND METHODS

### Study sites

This study recruited patients over a period between June 2020 and February 2022 from Aga Khan University Hospital, Nairobi, an urban metropolitan academic medical center. COVID-19 patients were recruited at hospitalization, upon confirmation of SARS-CoV-2 infection by RT-PCR. A few of these patients who presented to the hospital for non-COVID-19 treatments and had no COVID-19 symptoms, were also recruited as asymptomatic controls. We also recruited patients from Kilifi County Hospital, a community-based government hospital serving a rural coastal region. The Kilifi COVID-19 patients comprised mainly of outpatients with mild COVID-19 symptoms and their social contacts with asymptomatic SARS-CoV-2 infections, identified through COVID-19 surveillance.

### Ethical Approvals

This study received approval from the Kenya Medical Research Institute’s Scientific and Ethics Review Unit (KEMRI SERU; protocol no. 4081), and the Aga Khan University, Nairobi, Institutional Ethics Review Committee (protocol no. 2020 IERC-135 V2).

### Study design

This is part of an ongoing longitudinal cohort study that aimed at understanding SARS-CoV-2 immune responses among COVID-19 patients. The recruitment excluded plasma samples obtained after COVID-19 vaccination. After offering informed consent and having been confirmed COVID-19 positive with RT-PCR SARS-CoV-2 testing, the recruited patients had a blood sample taken which formed the sampling baseline timepoint hereafter referred to as Day 0. Follow up blood samples were taken on Days 7, 14, 28 and 180 (6 months) of a positive COVID-19 test. Plasma was immediately isolated from blood and stored at -80^°^C until the time of antibody measurements. Clinical data from these patients was entered and managed in Redcap.

### Grading COVID-19 severity

Grading of COVID-19 presentations was done according to the National institute of health (NIH) guidelines (42). “Asymptomatic” patients were positive for RT-PCR SARS-CoV-2 but were without COVID-19 symptoms. Patients were classified as “mild” if they were positive for SARS-CoV-2, and presented with fever, sore throat, cough, malaise, headache, muscle pain, vomiting, nausea, diarrhea, anosmia and ageusia. However, mild patients were devoid of shortness of breath, dyspnea, or abnormal chest imaging. Patients were classified as “moderate” if they were positive for SARS-CoV-2 RT PCR and exhibited proof of infection of the lower respiratory tract either via clinical examination, or through imaging. Oxygen saturation - (SpO_2_) of ≥94% on room air was also a characteristic of this moderate group. “Severely” sick patients were positive for SARS-CoV-2 RT PCR and were characterized by SpO_2_ levels of <94% on room air, a ratio of arterial partial pressure of oxygen to fraction of inspired oxygen (PaO_2_/FiO_2_) <300 mm Hg, breaths/minute of >30 breaths/min, and/or infiltration of the lungs of >50%. “Critically ill” patients were positive for SARS-CoV-2 RT PCR, and had evidence of respiratory failure, septic shock and/or multi-organ dysfunction syndrome.

### Anti-spike IgG antibody ELISA

Anti-spike IgG antibodies were measured using a previously described in-house ELISA targeting the full-length trimeric spike (35). The sensitivity of the ELISA was 0.927 (95% CI 0.879–0.961) estimated in 174 PCR positive Kenyan adults and a panel of sera from the UK National Institute of Biological Standards and Control (NIBSC) (35). The specificity was 0.99 (95% CI 0.981– 0.995) estimated in 910 sera collected from individuals in Kilifi in 2018 before the pandemic (35). Positive and negative controls were included in all the ELISA runs and results from these were reproducible (35)(43). A WHO international standard (44), calibrated at a maximum arbitrary value of 1000 binding antibody units per ml (BAU/ml) was included in the runs and used to quantify the antibody levels. Therefore, anti-spike IgG data were presented as BAU/ml based on that standard.

### Anti-RBD (IgA, IgG, IgM) ELISA

We used a commercial RBD ELISA kit (Mabtech AB, Nacka strand, Sweden) (Lot number 3890-1H-R1-1) which detects RBD-spike binding Igs of the isotypes (IgA, IgM, and IgG) with a specificity: negative percent agreement (NPA) of 100% (51/51 un-infected controls) and a sensitivity: positive percent agreement (PPA) of 100% (72/72 COVID-19 convalescents - PCR confirmed) (45). We optimized the kit for sample dilution through serial dilutions of randomly picked plasma samples from the 585 patients plasma samples from COVID-19 patients and available controls, taken prior to the COVID pandemic and available at the KEMRI Wellcome Trust Research Program (KWTRP) Biobank. 96-well plates were coated overnight at 4^°^C with 100μl per/well of strep-tactin® XT diluted at 1μg/ml. Coating solution was then discarded, and blocking done using 200μl of the dilution buffer per well for 1 hour. Washing was then done 5 times using 300μl/per-well of wash buffer. Plasma was diluted at 1:64 using the dilution buffer and 50μl was added in duplicate to the plates followed by 50μl/per-well of RBD Bridge reconstituted at 1:12. The plates were then placed on an orbital plate shaker at 400 revolutions per minute for 2 hours at room temperature (RT). The plates were then washed 5 times. Anti-WASP-HRP was diluted 200 times and was added at 100μl/per-well to the plates and incubation done for 1 hour at RT. Washing was then done 10 times and 100μl/per well of Tetra-methyl benzidine (TMB) was added. Plates were then incubated for 15 mins in the dark at RT before adding 100μl/per-well of stop solution. Absorbance was measured at 450nm using Bio Tek Synergy H1 microplate reader. To quantify the antibody levels, hyper immune plasma with a top standard dilution at 1:650, was assigned an arbitrary 1000 ELISA units and diluted serially, 2-fold. Therefore, anti-RBD Igs data were presented as arbitrary ELISA units (AEU). A 4-parameter logistic model generated by Gene5™ analytical software was used to determine the arbitrary concentrations of the antibodies. 10 pre-pandemic negative control plasma samples were included in the assay. These negative control plasma samples were used to establish a cutoff of sero-positivity calculated at 6 standard deviations of the mean of negative controls, according to the manufacturer’s instructions.

### SARS CoV-2 pseudo-virus production and Neutralization assay

Pseudo viruses were generated by co-transfecting MLV-gag/pol, MLV-CMV-Luciferase and SARS-CoV-2 Wuhan-1 spike plasmids with PEIMAX (Polysciences) into HEK293T/17 cells. Culture supernatants were clarified of cells (after 72 hours) by 0.45μM filter and stored at -70°C. HeLa-hACE2 cells, modified to overexpress human ACE2, were cultured in DMEM with GlutaMAX (Gibco-BRL) containing 10% heat-inactivated HIFCS and 1% of 100x Pen Strep at 37°C, 5% CO_2_. At confluency, cell monolayers were disrupted using 0.25% trypsin-1 mM disodium EDTA (Gibco-BRL). Plasma samples were heat-inactivated, clarified by centrifugation and serially diluted in 96-well flat-bottom culture plates (Corning, 3595) containing growth medium. Pseudo viruses and serially diluted plasma were incubated for 1 hour at 37°C, 5% CO_2_. HeLa-hACE2 cells were added at 10,000 cells per well and incubated for 72 hours at 37°C, 5% CO_2_. Supernatants were removed, the cells were lysed in Glo lysis buffer 1X (Promega, E2661), luciferase activity was measured by adding Bright-Glo (Promega, E2650) and luminescence was measured using Biotek Synergy H1 plate reader. Neutralization was measured as described by a reduction in luciferase gene expression after single round infection of Hela-hACE2 cells with spike-pseudo typed viruses. Levels were calculated as the reciprocal plasma inhibitory dilution (ID_50_) causing 50% reduction in relative light units (RLU).

### Statistics

Comparisons of antibody levels from one time point to the next were done using Friedman’s test of repeated measures with Pairwise Wilcoxon signed-rank tests for multiple comparison. Lowess regression curves were generated to indicate the overall antibody trajectories overtime. Spearman’s correlation coefficients were utilized to correlate RBD Igs, anti-spike IgG and neutralization ID_50_ antibody levels with each other, at each time point. Associations of disease severity, effect of change in time, age, and gender on antibody responses, were estimated using a linear mixed model. Kruskal Walli’s test with Dunn’s multiple comparison test was used to compare antibody levels across clinical severities. Mann-Whitney U test was used to perform non-parametric pairwise comparisons. Analyses were performed using R software V4.1.2 (46) and GraphPad prism V8.0.2. A P value of < 0.05 was considered statistically significant.

## RESULTS

### COVID-19 Cohort Recruitment

We had recruited a total of 309 COVID-19 patients, from whom a total of 585 blood samples had been collected across the 5 time points described here. From AKUH, Nairobi, we recruited a total of 270 hospitalized COVID-19 patients between June 2020 to September 2021, and from KCH, Kilifi, we recruited 39 COVID-19 patients between March 2021 to February 2022. Not every patient was available for all their scheduled times for follow-up samples. Thus, whilst 49.5% (153/309) of the patients had samples available from multiple time points, the rest only contributed to samples at Day 0 only (**Fig. 1**).

**Figure 1:**
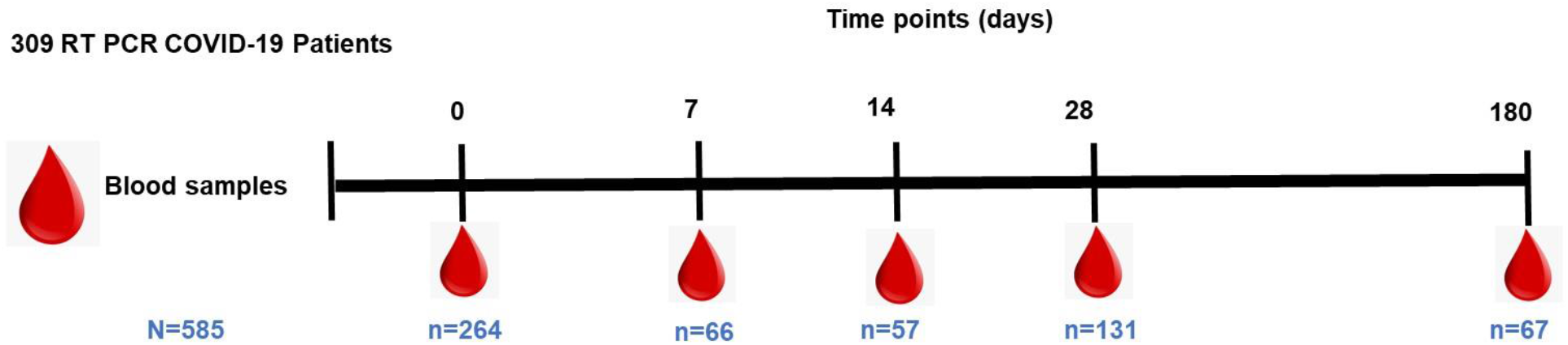
Longitudinal blood sampling over 6 months of COVID-19 diagnosis. The total number of plasma samples collected at each timepoint are indicated.

### Clinical characterization of the COVID-19 cohort

70% of the patients were male, and the median ages increased with disease severity (**Table 1**). Mild and severe groups had the most patients with 39.2% (121/309) and 45.6% (141/309), respectively, of the total number of patients. The rest comprised of asymptomatic at 6.8% (21/309), moderate at 7.4% (23/309), and critical at 0.9% (3/309) groups. Underlying non-communicable diseases were prevalent, especially among the severe and critically ill patients.

**Table1:** Patient’s baseline characteristics categorized according to the different grades of COVID-19 severities. Table generated using “Tableone” package in R. The continuous non-parametric variable is represented as median with inter-quartile range (IQR). Categorical variables are represented as frequencies.

|  | Asymptomatic | Mild | Moderate | Severe | Critical |
| --- | --- | --- | --- | --- | --- |
| n | 21 | 121 | 23 | 141 | 3 |
| **Demographics** |  |  |  |  |  |
| Age (median [IQR]) | 35.00 [29.00, 54.00] | 46.29 [36.22, 54.47] | 46.00 [35.50, 56.89] | 52.21 [43.00, 59.48] | 67.15 [60.06, 69.14] |
| Gender = Male (%) | 12 ( 57.1) | 77 ( 63.6) | 16 ( 69.6) | 108 ( 76.6) | 2 ( 66.7) |
| = Female (%) | 9 (42.9) | 44 (36.4) | 5 (30.4) | 33 ( 23.4) | 1 (33.3) |
| Race (%) |  |  |  |  |  |
| African American / Black | 21 (100.0) | 106 ( 87.6) | 20 ( 87.0) | 125 ( 88.7) | 3 (100.0) |
| Asian | 0 ( 0.0) | 6 ( 5.0) | 2 ( 8.7) | 5 ( 3.5) | 0 ( 0.0) |
| Caucasian | 0 ( 0.0) | 5 ( 4.1) | 1 ( 4.3) | 4 ( 2.8) | 0 ( 0.0) |
| Indian | 0 ( 0.0) | 4 ( 3.3) | 0 ( 0.0) | 7 ( 5.0) | 0 ( 0.0) |
| **Comorbidities** |  |  |  |  |  |
| Diabetes = Yes (%) | 0 ( 0.0) | 45 ( 37.2) | 6 ( 26.1) | 79 ( 56.0) | 3 (100.0) |
| Hypertension = Yes (%) | 0 ( 0.0) | 27 ( 22.3) | 3 ( 13.0) | 46 ( 32.6) | 3 (100.0) |
| HIV Positive = Yes (%) | 0 ( 0.0) | 5 ( 4.1) | 0 ( 0.0) | 5 ( 3.5) | 1 ( 33.3) |
| Renal disease = Yes (%) | 0 ( 0.0) | 1 ( 0.8) | 0 ( 0.0) | 3 ( 2.1) | 0 ( 0.0) |
| COPD = Yes (%) | 0 ( 0.0) | 0 ( 0.0) | 0 ( 0.0) | 1 ( 0.7) | 0 ( 0.0) |
| Cancer = Yes (%) | 0 ( 0.0) | 0 ( 0.0) | 0 ( 0.0) | 2 ( 1.4) | 0 ( 0.0) |
| **Patient management (%)** |  |  |  |  |  |
| Hospital admission= Yes (%) | 0 ( 0.0) | 103 ( 85.1) | 23 (100.0) | 141 (100.0) | 3 (100.0) |
| In ICU = Yes (%) | 0 ( 0.0) | 0 ( 0.0) | 1 ( 4.3) | 23 ( 16.3) | 3 (100.0) |
| **Outcome** |  |  |  |  |  |
| Outcome = Died (%) | 0 ( 0.0) | 1 ( 0.83) | 1 ( 4.3) | 6 ( 4.3) | 0 ( 0.0) |

### Anti-SARS-CoV-2 binding antibody sero-conversion rates and maintenance for the first 6 months of positive COVID-19 diagnosis

The levels of anti-spike IgG and anti-RBD Igs binding antibodies were strongly correlated with each other at all the time points: Day 0 (r =0.8385, P<0.0001), Day 7 (r =0.851, P<0.0001), Day 14 (r =0.841, P<0.0001), Day 28 (r =0.824, P<0.0001) and Day 180 (r =0.8379, P<0.0001), Spearman’s correlation coefficient) (**Fig. 2A**).

**Figure 2:**
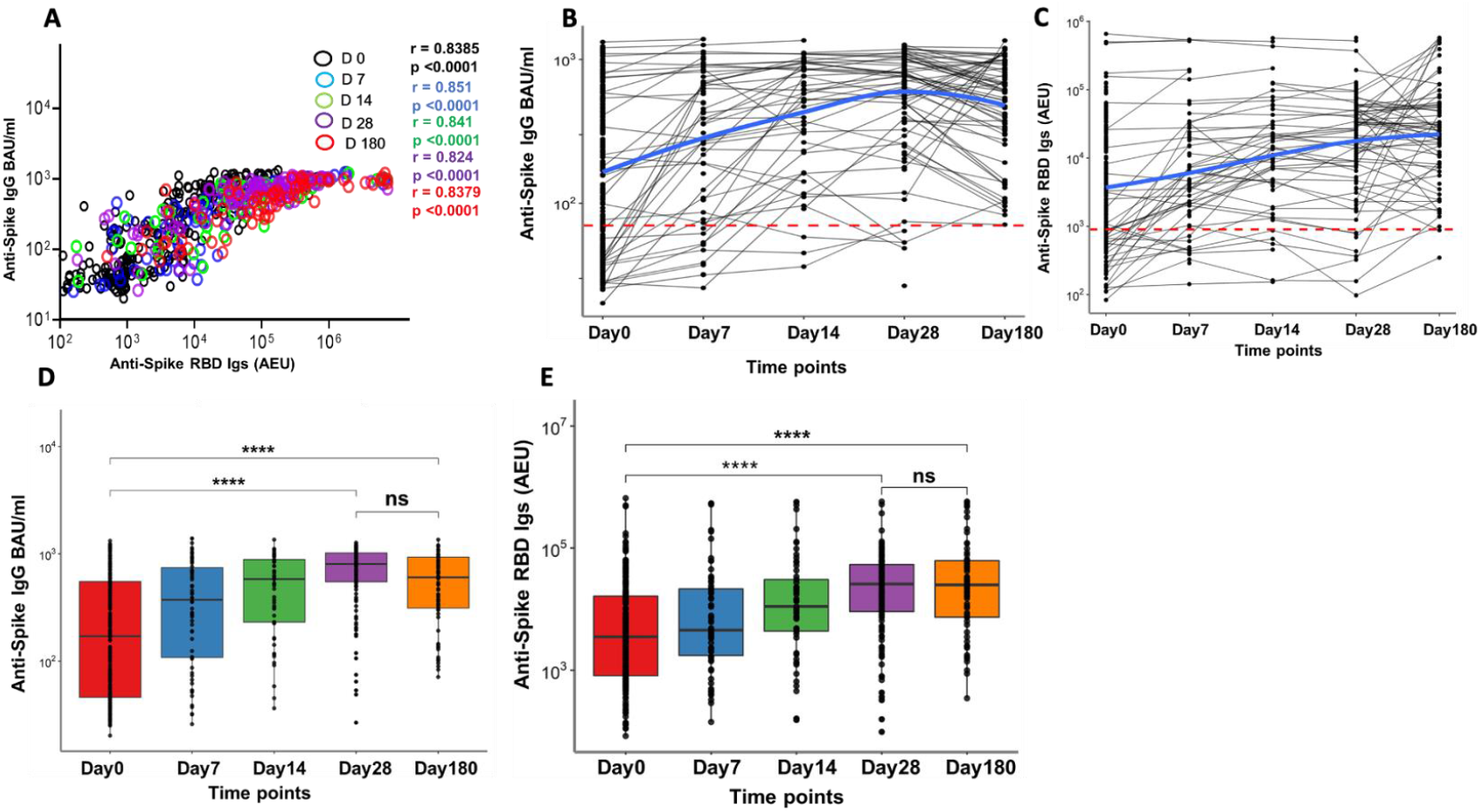
Antibody level kinetics, and correlation of anti-spike IgG and anti-spike-RBD immunoglobulins. (A) Schematic depicting a Spearman’s correlation of anti-Spike IgG binding antibody units (BAU) and anti-spike-RBD Igs arbitrary ELISA unit (AEU) levels at all the time points. (B) Kinetics of anti-spike IgG and (C) anti-spike-RBD Igs over the first six months of COVID-19 diagnosis. The dotted red line represents the threshold of sero-positivity defined by the negative controls as described in the methods. The blue curve depicts the best fit non-linear regression model of antibody trajectories. (D) Box plots depicting anti-spike IgG antibody kinetics and anti-spike-RBD Igs (E) over 6 months as calculated with the Friedman’s test of repeated measures with Pairwise Wilcoxon signed-rank test for multiple comparison correction. *P < 0.05, **P < 0.01, ***P < 0.001, ****P < 0.0001

The rates of sero-conversion increased with time of COVID-19 diagnosis (**Fig. 2 B and C**). Of the patients with plasma samples from more than two time points, 7.8% (12/155) had antibody levels increasing sharply from Day 28 to Day 180, suggesting possible re-infections. Only 63.6% (168/264) of the patients had sero-converted for spike IgG at Day 0, whilst all the patients whose samples were available at Day 180 were antibody positive (67/67). When anti-spike RBD Igs were considered, 69.3% (183/264) of the patients had sero-converted at Day 0, and of these, 97% (65/67) of those who had multiple successive samples available remained antibody positive at Day 180 (**Table 2**).

**Table 2:** SARS-CoV-2 sero-conversion rates. Percentages seropositivity and sero-negativity overtime. Sero-conversion cutoffs were determined using the negative controls as described in the methods.

| Anti-spike IgG |  |  |  |
| --- | --- | --- | --- |
| Time point | Plasma samples | Sero-positivity = n (%) | Sero-negativity= n (%) |
| Day 0 | 264 | 168 (63.6) | 96 (36.4) |
| Day 7 | 66 | 54 (81.8) | 12 (18.2) |
| Day 14 | 57 | 54 (94.7) | 3 (5.3) |
| Day 28 | 131 | 126 (96.2) | 5 (3.8) |
| Day 180 | 67 | 67 (100) | 0(0.0) |

| Anti-spike-RBD Igs |  |  |  |
| --- | --- | --- | --- |
| Time point | Plasma samples | Sero-positivity = n (%) | Sero-negativity = n (%) |
| Day 0 | 264 | 183 (69.3) | 81 (30.7) |
| Day 7 | 66 | 55 (83.3) | 11 (16.7) |
| Day 14 | 57 | 51 (89.5) | 6 (10.5) |
| Day 28 | 131 | 121 (92.4) | 10 (7.6) |
| Day 180 | 67 | 65 (97.0) | 2(3.0) |

Anti-spike IgG levels increased from a median level of 171.6 BAU/ml (IQR, 46.00 - 553.8) from Day 0 to 804.57 BAU/ml (IQR,550.37 - 1021.74) at Day 28 (P<0.0001, Friedman test with Pairwise Wilcoxon test for multiple comparison). Anti-spike-RBD Igs increased from a median of 3510.38 AEU (IQR, 811.73 -16111.00) at Day 0 to 25723.71 AEU (IQR, 9098.85 - 53537.56) at Day 28 (P<0.0001) (**Fig. 2 D and E**). Antibody levels were thereafter maintained without significant decay with Day 180 median levels for anti-spike IgG being 604.42 BAU/ml (IQR,312.58 - 933.05) and for anti-spike-RBD Igs being 25002.75 AEU (IQR,7343.15 - 62111.00), which were like the respective levels for day 28 in both measurements (p >0.9). Overall, anti-spike IgG antibody BAU/ml levels increased with time of COVID-19 diagnosis (Estimate = 0.36, P<0.0001), after adjusting for gender and age, in a linear mixed model analysis of patients with at least more than two plasma samples (**Supplementary table 1**).

### Kinetics of neutralization antibody levels over the first 6 months of COVID-19 diagnosis

We further determined if the antibodies induced by SARS-CoV-2 infection could elicit viral neutralization in a subset of 51 patients for whom plasma samples were available at days 0, 28, and 180 of infection. These 51 patients comprised of 7 asymptomatic, 21 mild, 21 severe, 1 moderate and 1 critically sick. Due to the single participant in both moderate and critical ill categories, we included the moderately sick patient in the mild group (with whom the clinical presentation was similar), while the critically sick patient was included in the severe group to power the statistical comparison. These 51 individuals had similar antibody kinetics to what was observed for the entire study as reported above and therefore representative of the study population (**Supplementary figure 1**).

A total of 90.2% (46/51) of the patients had already developed neutralizing antibodies by Day 0. At Day 28, the proportion of responders had increased to 96.1% (49/51), and it was 94 % (48/51) at Day 180. For all the participants, antibody neutralization potency increased significantly from a median 50% inhibitory dilution (ID_50_) of 155.10 (IQR, 42.92 - 578.79) at Day 0 and peaked at Day 28 post infection at a median ID_50_ of 621.88 (IQR, 145.89 - 1958.61) (p<0.0001, Friedman test with Pairwise Wilcoxon for multiple comparison), before rapidly declining to a median ID_50_ of 153.00 (IQR, 66.07-326.08) at Day 180 (p= p<0.0001,, Friedman test with Pairwise Wilcoxon for multiple comparison) (**Fig. 3A**). To quantify the decline, we fitted a linear mixed model describing the kinetics of neutralization antibody levels (**Supplementary table 2**), and then extracted the fitted values to obtain independent estimates of neutralizing antibody levels at each timepoint. When the independent estimate fitted values of ID_50_ 1929.928 at Day 28 were compared with those of ID_50_ 630.9376 at Day 180, we found that the neutralizing antibody levels had decreased by three-fold (**Fig. 3B**).

**Figure 3:**
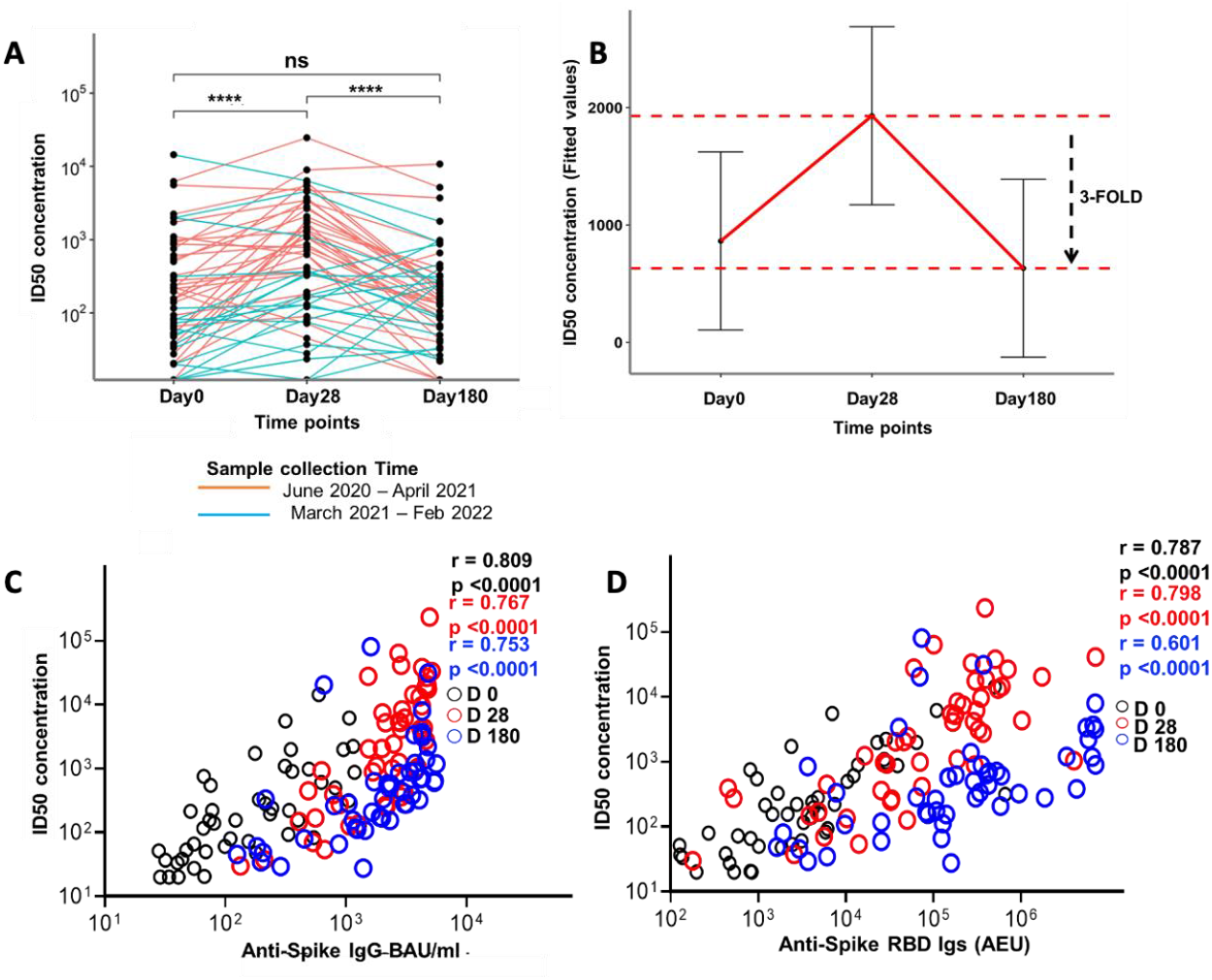
Neutralizing ID50 potencies of plasma-antibodies against a Wuhan-based SARS-CoV-2 pseudo virus, their correlations with binding antibodies over time and kinetics of neutralizing ID50. (A) Kinetics of neutralizing ID50 potencies over three time points of COVID-19 diagnosis colored according to the plasma sample collection time. Statistics were done with the Friedman’s test for repeated measures with Pairwise Wilcoxon signed-rank test for multiple comparison correction. (B) Fitted values from a linear mixed model to depict rate of decline in neutralizing antibodies towards Day 180. (C) Spearman’s correlation between neutralizing ID50 and anti-spike IgG and (D) between neutralizing ID50 and anti-spike-RBD Igs at the three-time points *P < 0.05, **P < 0.01, ***P < 0.001, ****P < 0.0001.

Interestingly, for those participants who had been recruited earlier in the pandemic (pre-April 2021), the trajectory of antibody neutralization levels depicted an increase towards Day 28 post infection followed by a decline towards Day 180 (**Fig. 3A**). However, for those recruited later in the pandemic, (post-April 2021), the responses were more heterogenous, with neutralizing antibody levels increasing over time and plateauing between Day 28 and Day 180.

To establish if the magnitude of the neutralization antibody potencies varied with changes in the circulating SARS-CoV-2 strains at the time of recruitment, we categorized participants based on available data in the public domain on the SARS-CoV-2 strains that were circulating in Kenya circulating at the time of patient sampling, as we did not have the sequence data to a certain the precise strain infecting the patients. On this basis, participants were categorized into two groups, those recruited earlier in the pandemic and hence likely to have been infected with the Wuhan strain, and those recruited later, and hence likely to have been infected by other SARS-CoV-2 variants like alpha, beta, delta and omicron (47)(48).

We observed higher neutralizing antibody potency from the Day 0 samples of those recruited before April 2021 with a median ID_50_ of 222.06 (IQR, 92.73-874.11) as compared to those sampled after April 2021 with a median ID_50_ of 49.47 (IQR, 20.00-82.79) (P<0.01) (**Supplementary figure 2**). At day 28, pre-April 2021 plasma samples maintained higher neutralizing antibody potency with median ID_50_ 1287.73 (521.54 - 3059.15) as compared to post-April 2021 plasma samples ID_50_ 129.83 (IQR, 56.04 - 356.96) (P<0.0001, Mann Whitney test). At day 180, both groups had similar neutralizing antibodies potencies at median ID_50_ of 2021 151.02 (IQR, 41.55 – 218.77), pre-April 2021 168.93 (IQR, 75.56 – 408.75) post-April 2021 (p=0.38, Mann Whitney test).

When antibody neutralization potencies were correlated with the binding antibody levels, high correlations were observed: for the IgG antibodies against the spike protein, r =0.809 (P<0.0001, Spearman’s correlation coefficient) at Day 0, r=0.767 (P<0.0001) at Day 28, and r =0.753 (P<0.0001) at day 180 (**Fig. 3C**), and for the spike-RBD Igs, r =0.787 (P<0.0001) at Day 0, r =0.798 (P<0.0001) at Day 28, and r =0.601 (P<0.0001) at Day 180 (**Fig. 3D)**.

### Associations of spike binding and virus neutralizing antibody potencies with COVID-19 severity

Increasing levels of binding antibodies were positively associated with disease severity with asymptomatic and severe patients having the lowest, and the highest antibody levels respectively, at earlier time points of the infection (**Fig. 4A**). For instance, Day 0 anti-spike IgG levels were higher among the severe than the mild group, with median levels of 250.90 (IQR, 64.37 - 700.00) and 115.20 (IQR, 40.41 - 406.5) BAU/ml, respectively (p = 0.018, Kruskal Wallis test with Dunn’s test for multiple comparison). Similarly, Day 28 anti-spike IgG antibody levels were higher among the severe than the asymptomatic group, with median levels of 855.80 (IQR, 615.00 - 1099.00) and 189.10 (IQR, 69.23 - 559.60) BAU/ml, respectively (P<0.0001). Day 28 anti-spike IgG antibodies were also higher among the mild than the asymptomatic, with median levels of 821.10 (IQR, 407.50 - 1024) and 189.10 (IQR, 69.23 - 559.60), respectively (P<0.0001). The moderate group had higher anti-spike IgG antibody levels than the asymptomatic group, with median levels of 797.7 (IQR, 690.80 - 916.2) and 189.10 (IQR, 69.23 - 559.60) BAU/ml, respectively (p=0.03). This positive association between increasing anti-IgG levels and disease severity remained significant in a linear mixed model, even after adjusting for gender and age (Estimate= 0.184741, P<0.01, **Supplementary Table 1**). However, at Day 180, these antibodies were all similar in all the pairwise group comparisons.

**Figure 4:**
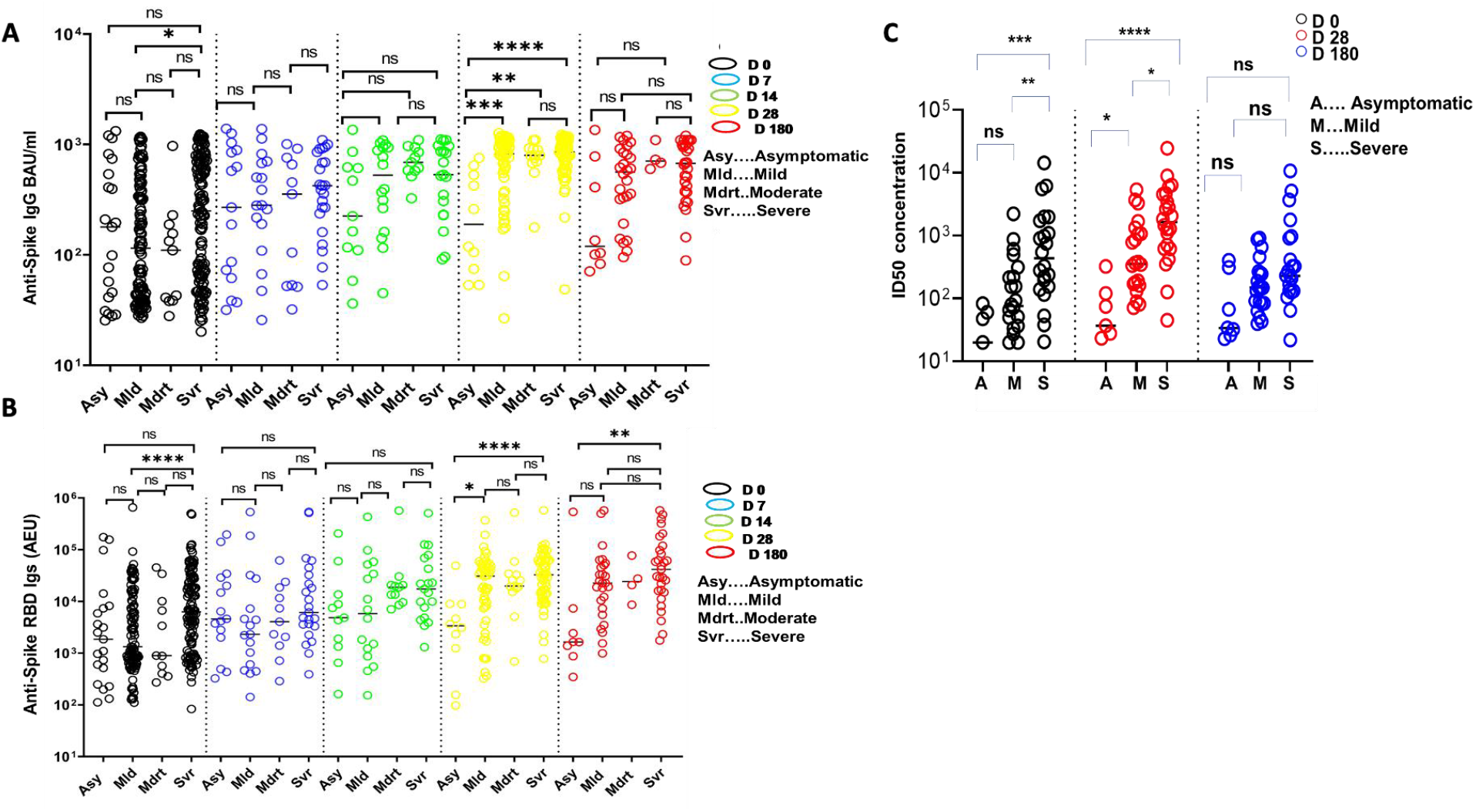
Comparisons of antibody levels and SARS-CoV2 viral loads (Ct values) across different grades of COVID-19 severities; Schematic depicting comparisons of antibody levels among clinical groups, anti-spike IgG (A) and anti-spike-RBD Igs (B). (C) A comparison of neutralizing ID50 potencies across disease phenotypes at each of the three time points. All comparisons were done using Kruskal-Walli’s method with Dunn’s multiple comparison correction. * P < 0.05, **P < 0.01, ***P < 0.001, ****P < 0.0001.

Day 0 neutralizing antibody potencies were higher among severe than mild (P=0.01) and asymptomatic individuals (P<0.001), with medians ID_50_ being 436.50 (IQR, 151.70 - 1795.00) 76.04 (IQR, 31.96 - 244.90) and 20.00 (IQR, 0.00 - 60.64), respectively (**Fig. 4C**). At Day 28, severe patients maintained higher neutralizing antibody potencies with a median ID_50_ of 1653 (IQR, 621.70 - 4589.00) than the asymptomatic ID_50_ 37.03 (IQR, 23.44 - 121.10) (P<0.0001), and mild patients median ID_50_ 357.00 (IQR, 153.90 - 1138.00) (p=0.03). The mild patients had higher neutralizing antibody potencies than asymptomatic individuals (p= 0.04). However, the neutralization antibody potencies had dropped significantly by Day 180 and were no longer different in any of the pairwise group comparisons.

### Associations of viral loads with COVID-19 severity

Because differences in viral loads could explain the associations between antibody levels and functions with disease severity, we compared viral loads among groups using cycle threshold (Ct) values from the Day 0 RT-PCR SARS-CoV-2 tests where data were available. The comparisons within each of the AKUH and KCH hospitals were done separately as different RT-PCR assays had been used. This was done for a total of 189 patients (asymptomatic =13, mild=67, moderate= 17, severe= 92), for whom viral-load data were available. We found no evidence that the viral loads were associated with the different clinical phenotypes (**Supplementary figure 3**). However, we recognize the limitation of small numbers in the groups.

## DISCUSSION

Most of the COVID-19 patients in this study sero-converted and maintained SARS-CoV-2 spike binding antibodies for the first 6 months after positive COVID-19 diagnosis, without significant decay. In a subset of the individuals, antibodies with virus neutralizing function against a Wuhan SARS-CoV-2 strain-based pseudo virus were observed, which peaked 28 days of COVID-19 diagnosis. However, neutralizing antibody potency had dropped three-fold by six months of COVID-19 diagnosis. Furthermore, the ability to neutralize the pseudo virus was higher in plasma samples collected during the period when the Wuhan strain was spreading and then diminished, but not completely abolished, in the collected later in the pandemic when the subsequent SARS-CoV-2 variants of concern were the most prevalent strains. Both the antibody-binding levels and neutralizing potencies were strongly correlated and increased with disease severity.

The observation that binding antibody levels were stable for the first six months of COVID-19 diagnosis, is consistent with some of the previous reports in populations from high income countries (HIC), where anti-SARS-CoV-2 antibodies were maintained for the first few months of infection (21,22,49–51). However, other studies from HIC populations found binding antibodies to wane quickly within the first few months after infection (17,52–54). Very few studies have determined the kinetics of anti-SARS-CoV-2 antibody kinetics in Sub Saharan African populations. A study from Ethiopia followed PCR confirmed COVID-19 patients for a median time of 31 days and collected blood samples every 3 days during follow-up. They used lateral flow immunoassays to quantify anti-spike and anti-nucleoprotein IgM, IgG, and an electro-chemiluminescent assay to quantify total anti-nucleoprotein immunoglobulins. They reported heterogenous antibody kinetics and a lack of sero-conversion among some patients (27). A South African study reported rapid waning of anti-SARS-CoV-2 RBD IgA and IgM, but a better maintenance of IgG antibodies quantified weekly over a period of 3 months among COVID-19 patients (55). Our results suggest a longer maintenance of anti-SARS-CoV-2 antibodies and highlight the importance of longer durations of follow-ups in longitudinal studies ascertaining antibody kinetics following infection.

Our finding, that neutralizing antibodies decayed three-fold by the sixth month of follow-up, suggests that anti-SARS-CoV-2 antibodies with neutralizing function are short-lived. This finding is in agreement with previous studies in the UK, Canada, and USA, which also found neutralizing antibodies to last for a few months (18,19,50,54). Only very few studies have reported anti-SARS-CoV-2 neutralizing antibodies to persist for more than six months (21,56), which is concerning given that neutralization is one of the most important immune mechanisms against viruses. This probably explains possible re-infections in our study, where a few patients had binding and neutralizing antibodies increasing after Day 28 of COVID-19 diagnosis (without vaccination). In agreement, occurrences of SARS-CoV-2 re-infections have also been reported from other studies(57)(58).

The decreasing ability for plasma antibody to neutralize the Wuhan-based strain pseudo virus with the time of sampling probably reflects the subsequent gradual replacement of the Wuhan strain with others like alpha, beta, delta and omicron in Kenya (47)(48). Better neutralization potencies to the infecting strain, and reduced neutralization to alternative stains has also been observed in other studies. For example, recognitions of B.1.1.7-Alpha and B.1.351-Beta strains were reduced in antibodies elicited by the parental Wuhan and D614G strains (59–61).

Nonetheless, the fact that we could still detect antibodies with neutralization potency against the Wuhan strain-based pseudo virus throughout the six months of follow up in this longitudinal study indicates a good level of cross reactivity across the different variants as reported elsewhere (62). Further assessment of cross-neutralization among different variants will be important in understanding the extent of infection induced immunity in providing protection from future SARS-CoV-2 variants during periods of high transmission, as well as the effectiveness of the current SARS-CoV-2 vaccines. Therefore, it will be important to determine the neutralizing breath of antibodies generated against the parental Wuhan virus and COVID-19 immunizations, against upcoming SARS-CoV-2 variants of concern.

Both spike binding antibody levels and neutralization potencies increased with COVID-19 severity, and especially so in the first month of diagnosis. Similar findings were reported from Italy and USA where anti-spike and anti-spike-RBD antibody concentrations were found to be higher among severely than mildly sick COVID-19 patients during acute stages of infection (63)(64). Of note, a recent study on Kenyan COVID-19 patients, found symptomatic COVID-19 patients to have higher spike IgG antibodies as compared to asymptomatic patients (33). Neutralizing antibodies were also higher among severely than mildly sick USA COVID-19 patients (65), and ICU patients compared to non-ICU Chinese patients (66). Despite the clear finding of higher binding antibody levels and neutralizing function among the severe than mild and asymptomatic patients, the causal effect relationship is not clear. Higher viral loads, inflammation, or both, could explain the induction of higher antibody levels and neutralization potencies. However, our study did not find differences in viral loads among the disease severities, perhaps owing to the small sample in this study.

In conclusion, our results suggest that SARS-CoV-2 neutralizing antibodies are short-lived, whilst other antibodies may be long lasting. Hence, binding antibodies are not an accurate surrogate of neutralizing function, especially during convalescence as has also been reported elsewhere (51). Aside from neutralization, antibodies can also mediate viral clearance through other mechanisms via their fragment crystallizable (Fc) region, such as antibody dependent cellular cytotoxicity and complement deposition. However, such roles remain largely uninvestigated for SARS-CoV-2 (65). However virus neutralization is one of the main mechanisms by which COVID-19 vaccines work (67)(68), and may be more likely to offer sterile immunity compared to other antibody mediated mechanisms. Understanding antibody kinetics, both quantities and functions will be important in informing current vaccination strategies, and in the development of second generation COVID-19 vaccines that could offer sterile immunity, and the much-needed long term protection.

In the meantime, efforts to broaden and extend the current levels of predominantly naturally acquired anti-SARS-CoV-2 immunity in sub-Saharan African populations with vaccination, followed with regular administrations of regular vaccine boosters will be required to sustain the high levels of neutralizing antibodies that peak after acute infection.

## Data Availability

All data produced are available online at https://doi.org/10.7910/DVN/6KW9U1

## Abbreviations

AEU: Arbitrary ELISA units
BAU: Binding antibody units
COVID-19: Corona virus disease 2019
Ct: Cycle threshold
ELISA: Enzyme-linked immuno-sorbent assay
HRP: Horseradish peroxidase; Immunoglobulin G
IgM: Immunoglobulin M
IgA: Immunoglobulin A
Ig: Immunoglobulin
ID_50_: 50% inhibitory dilution
OD: Optical density
OPD: o-Phenylenediamine
PBS: Phosphate buffered saline; Room temperature
RT-PCR: Reverse transcription polymerase chain reaction
RBD: Receptor binding domain
RLUs: Relative light units
SARS-CoV-2: severe acute respiratory syndrome coronavirus 2
WASP-HRP: Wiskott-Aldrich syndrome protein - Horseradish peroxidase;

## Author’s contribution

FMN, EN and SS designed the study. FMN, EN, WUM, SS, RS and MS supervised the work and provided laboratory technical guidance. JNK, YS, PW and JM performed the laboratory technical work. JN, HK, BK, JG, DM, and GM assisted in laboratory antibody spike IgG analysis. KM assisted in statistical data analysis. AK, LM, VO, JS, JM, AM, and ZN assisted in data and sample collection as well as clinical data curation. PB, EN, AA, SS, RS, MS, IO and WUM offered guidance in developing the manuscript. JNK wrote the first manuscript draft. All authors contributed to writing and revising the final version of the manuscript.

## Availability of data and materials

The datasets and R/GraphPad Prism software codes used and/or analyzed in the current study are available from the corresponding author upon reasonable request.

## Consent for publication

Not applicable

## Acknowledgements

We appreciate all the study participants. We also thank the community field assistants and health care workers involved in the longitudinal blood sampling. Many thanks to the Initiative to Develop African Research Leaders (IDeAL) for the master’s sponsorship. We also sincerely appreciate Penny Moore, University of Witwatersrand for donating MLV-gag/pol, MLV-CMV-Luciferase and SARS-CoV-2 Wuhan-1 spike plasmids and Elise Landais, Deli Huang, and David Nemazee from The Scripps Research Institute for donating the HeLa-hACE2 cells.

