## Supplemental Data for "Kinetics of naturally induced binding and neutralizing anti-SARS-CoV-2 antibody levels and potencies among Kenyan patients with diverse grades of COVID-19 severity"

*Supplementary Material*

### Supplementary Figures and Tables

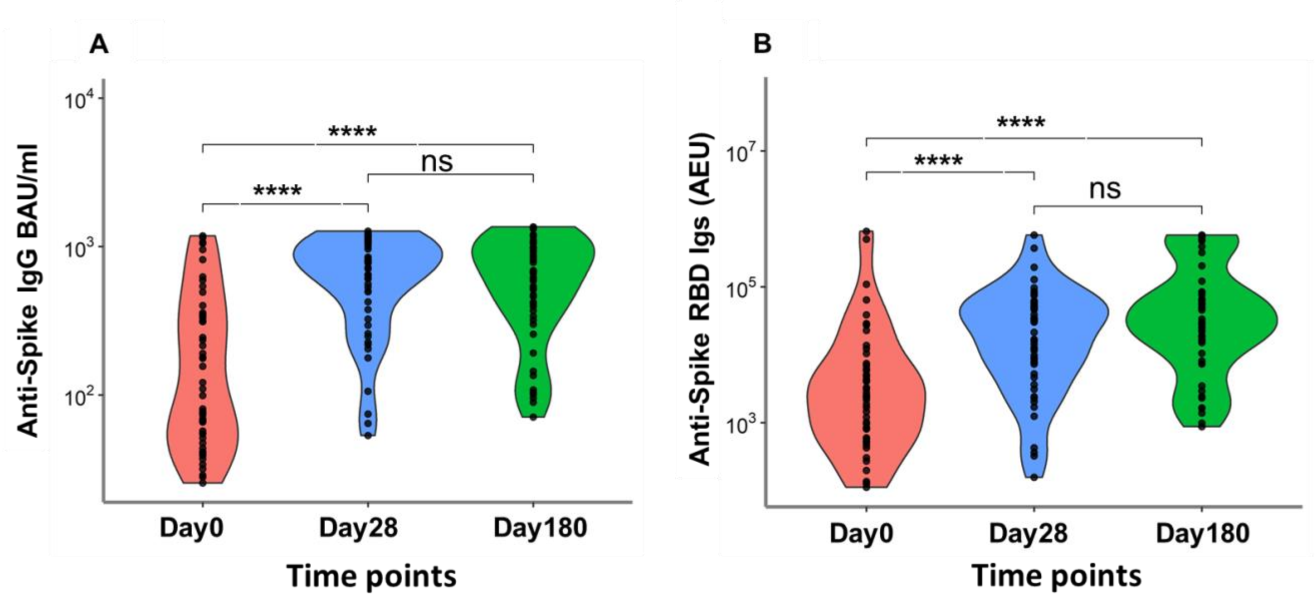


1. Supplementary Figure 1: Kinetics of antibody levels (A) Anti-spike IgG BAU/ml and (B)Anti-spike RBD Igs (AEU), in a subgroup of 51 COVID-19 patients. Subgroup was used in the neutralizing assay. Calculations were done with the Friedman's test for repeated measures with Pairwise Wilcoxon signed-rank test for multiple comparison correction. $P < 0.05$, $**P < 0.01$, $***P < 0.001$, $****P < 0.0001$.

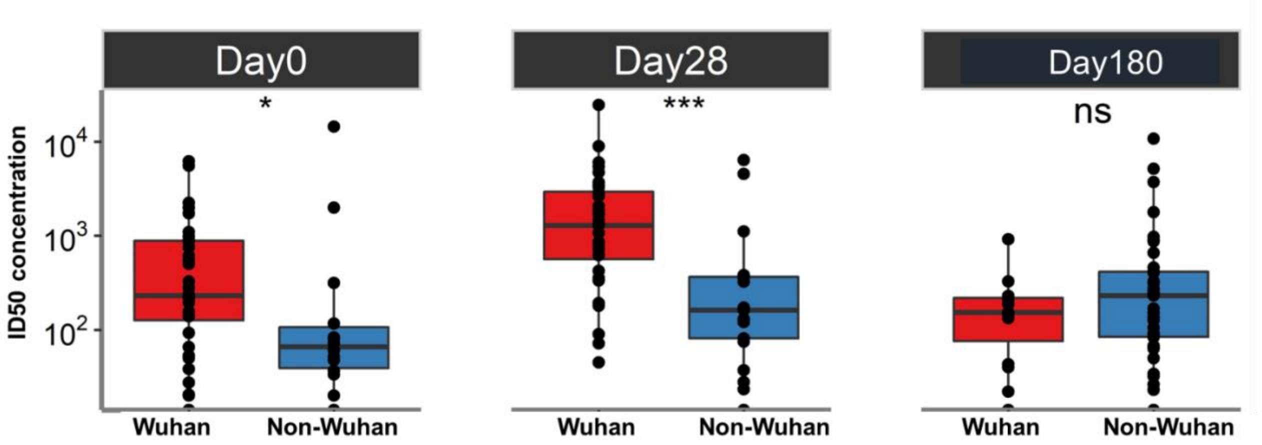


2. Supplementary Figure 2: Cross-reactive neutralization. Comparison of neutralizing $ID_{50}$ of samples collected early in the pandemic (pre-April 2021) during Wuhan strain prevalence and those collected later in the pandemic (post-April 2021) during the prevalence of other non-Wuhan strains such as alpha, beta, delta and omicron. Comparison's done using a Mann Whitney U test. $P < 0.05$, $**P < 0.01$, $***P < 0.001$, $****P < 0.0001$.

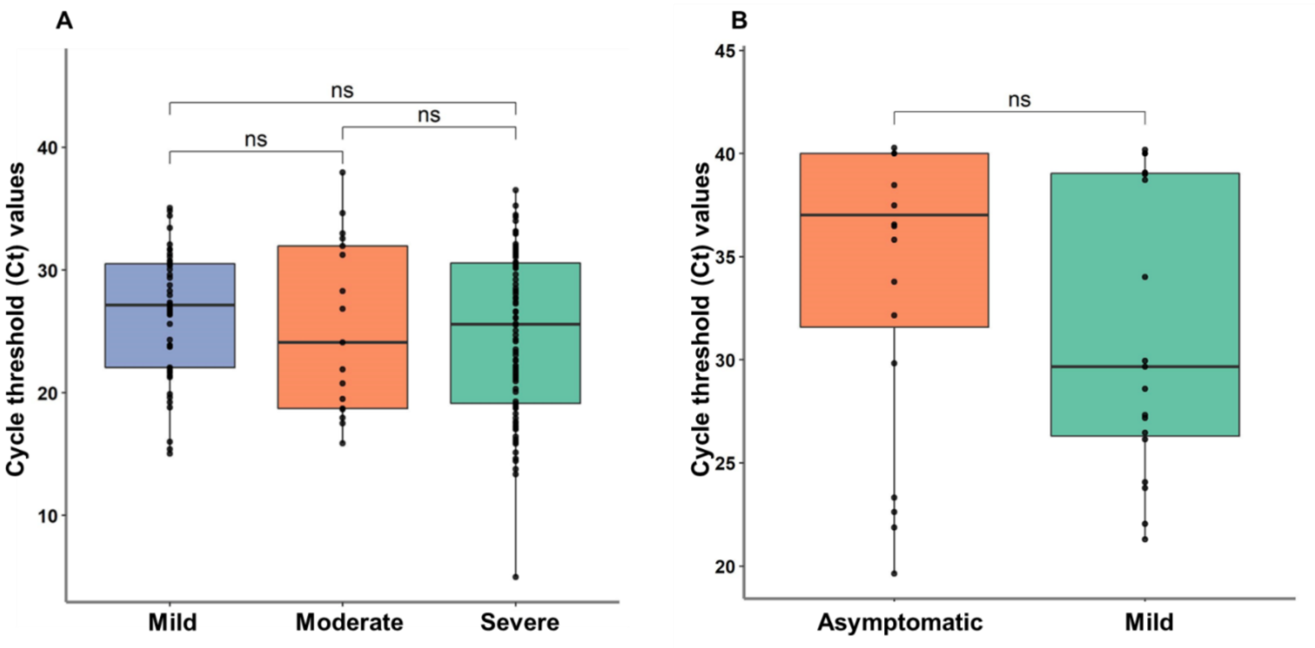


3. Supplementary Figure 3: Viral load comparisons among clinical groups. (A) RT PCR tests done in AKUH, Nairobi and (B) KCH, Kilifi . Calculated using Kruskal-Walli's test and Man Whitney U test respectively. *P* < 0.05, ***P* < 0.01, ****P* < 0.001, *****P* < 0.0001.

### Supplementary Tables

1. Supplementary Table 1: Linear mixed model outputs of association of anti-spike IgG antibodies levels with other variables. Captures patients with at least two plasma samples. Estimates indicates effect of change of the variables on measure of anti-spike IgG. The estimates are natural log transformed. Each estimate has a corresponding P value to indicate significance level.

| | *Estimate* | *Std. Error* | *Pr(>\|t\|)* | *Significance level* |
|---|---|---|---|---|
| *(Intercept)* | *3.901206* | *0.336231* | *<2e-16* | ***** |
| *Time* | *0.355905* | *0.026349* | *< 2e-16* | ***** |
| *Severity* | *0.184741* | *0.058897* | *0.00213* | **** |
| *Age* | *0.013027* | *0.005742* | *0.02491* | *** |
| *Gender* | *-0.146325* | *0.152025* | *0.33760* | *ns* |

*Signif. codes: 0 '***' 0.001 '**' 0.01 '*' 0.05 '.' 0.1 ' ' 1*

2. Supplementary Table 2: Modelling of neutralizing antibody kinetics. Estimates indicates effect of change of the variables on measure of neutralizing antibodies (Day 0 used as reference point). Fitted effects are obtained from a linear mixed model output. Each estimate has a corresponding P value to indicate significance level.

| | **Estimate (S.E)** | **Fitted Values (95% C.I)** | **p-value** |
|---|---|---|---|
| Day 0 | ref | 862.99 (104.9-1621.1) | |
| Day 28 | 1066.9 (403.4) | 1929.93 (1171.8-2688.1) | *0.00949* |
| Day 180 | -232.1 (403.4) | 630.94 (-127.1-1389.1) | *0.5664* |
